# Assessment of Corvis ST biomechanical parameters in diagnosing primary open-angle glaucoma in high myopia

**DOI:** 10.1101/2025.06.25.25330249

**Authors:** Bing Zhang, Yune Zhao

## Abstract

**PURPOSE:** The purpose of this study was to investigate the diagnostic value of the Corvis ST parameters in primary open-angle glaucoma (POAG) among high myopes.

**METHODS:** A total of 49 eyes (33 POAG cases and 16 controls) of 49 high myopes (axial length ≥26 mm) from open online datasets were analyzed. The Corvis ST parameters were compared between the POAG and the control eyes after adjusting for age and sex. Logistic regression models were used to analyze the associations between POAG and each Corvis ST parameter, with adjustments for age and sex. The receiver operating characteristic (ROC) curves were used to assess the diagnostic value of the significant parameters in the logistic models. The linear support vector machine (SVM) model was fitted using parameters demonstrating diagnostic values in the ROC analyses.

**RESULTS:** The means of the following biomechanical parameters were significantly lower in patients with POAG: A1 time (*P* = 0.004), A1 length (*P* = 0.011), A2 length (*P* = 0.035) and peak distance (PD, *P* < 0.001). Conversely, the mean A1 velocity (P < 0.001) and concavity curvature radius (*P* = 0.035) were significantly higher in eyes with POAG. In the logistic regression models, POAG was significantly associated with the A1 time (*P* = 0.014), A1 velocity (*P* = 0.003), A1 length (*P* = 0.043), and PD (*P* = 0.004). The ROC analyses demonstrated diagnostic utility for A1 time (area under the curve [AUC]: 0.77), A1 velocity (AUC: 0.78), and PD (AUC: 0.87). In the SVM model, the model demonstrated good performance, with a classification accuracy of 0.898.

**CONCLUSION:** In high myopia patients, Corvis ST parameters, including A1 time and PD, demonstrated a potential diagnostic value for POAG.

## Introduction

Diagnosis of glaucoma in high myopia can be a clinical challenge.^[1]^ On the one hand, the risk of glaucoma increases in myopic patients, with an estimated 20% higher odds per diopter, and the risk increases even more steeply in high-degree myopia.^[2,3]^ This indicates that it is more common to encounter glaucoma patients in high myopia patients. On the other hand, high myopia may complicate the diagnosis of glaucoma. Besides intraocular pressure (IOP), the morphology of the optic disc, the thickness of the retinal nerve fiber layer (RNFL), and the visual field are three important indicators of glaucoma,^[4]^ all of which may be atypical in patients with high myopia. First, in high myopia patients, the optic nerve changes may mimic glaucomatous findings, and interpreting the optic disc can be difficult.^[5]^ Second, both high myopia and glaucoma feature thinner RNFL thickness, and their alteration patterns may resemble each other.^[6]^ Third, the visual field defect in high myopia can mimic a glaucomatous defect in some patients, with one study reporting it in 16.1% of young Chinese high myopes.^[7]^ Therefore, it is important to find new biomarkers to support the diagnosis of glaucoma in high myopia. The association between corneal biomechanical parameters and the risk of glaucoma^[8]^ has been shown to be independent of IOP.^[9]^ In this study, we explored the diagnostic values of Corvis ST parameters in primary open-angle glaucoma (POAG) among high myopia, using logistic models, receiver operating characteristic (ROC) curves, and support vector machine (SVM) model.

## Methods

This study is based on the online open datasets provided by Asaoka *et al*. through CC BY license.^[10,11]^ Participants aged ≥20 years with axial length (AL) exceeding 26 mm were enrolled, while diabetics and contact lens wearers were excluded as corneal biomechanics may be affected. POAG was diagnosed clinically by experienced eye doctors. Participants with a history of other ocular diseases were excluded, except for those with clinically insignificant senile cataracts.^[10,11]^ The studies to generate the original datasets were approved by the Research Ethics Committee of the Graduate School of Medicine and Faculty of Medicine at The University of Tokyo, Saneikai Tsukazaki Hospital, and Hiroshima University Hospital and adhered to the tenets of the Declaration of Helsinki. Written informed consent was obtained from all participants.

Corvis ST (OCULUS Optikgeräte GmbH, Germany) measurements were performed three times per eye at a time interval of about 1 min, and the mean value of three measurements was analyzed. During each measurement, the cornea moves inward and then outward after the air-puff pulse, which generates two applanations (hereafter referred to as A1 and A2). The parameters from Corvis ST in this study are as follows: (1) A1/A2 time, the period from the release of the air puff to each applanation; (2) A1/A2 length, the length of the flattened cornea at each applanation; (3) A1/A2 velocity, the speed of the apex of the cornea at each application; (4) highest concavity time (HC time), the period taken to reach the maximum deformation; (5) deformation amplitude, the distance the apex of the cornea moves from the start to the HC; (6) peak distance (PD), the distance between the two peaks at the HC; (7) concavity curvature radius (CCR), the curvature radius at the apex during HC; and (8) central corneal thickness at rest^[10]^ The AL and mean corneal curvature were measured using IOL master 500 (Carl Zeiss Meditec., Germany). Goldmann applanation tonometry was carried out after topical anesthesia with fluorescein staining to obtain IOP.

The logistic regression models were used to explore the association between Corvis ST biomechanical parameters and POAG after adjusting for age and sex. For the Corvis ST parameters that were significantly associated with the presence of glaucoma, the ROC curve analyses were performed. Then, a supervised learning algorithm, the linear SVM, was used to build a diagnostic model using multiple parameters. The linear SVM analysis was performed using the SVM package from the Scikit-learn (Python 3.9). All the other analyses and the figure plotting were performed using STATA 18 (College Station, Texas, USA).

## Results

As shown in Table 1, 49 eyes of 49 participants (16 healthy controls and 33 patients with POAG) were enrolled for analysis. Regarding the Corvis ST biomechanical parameters, the mean A1 time in the whole sample was 7.23 ± 0.28 ms, and it was significantly lower in eyes with POAG (*P* = 0.004). Similarly, the mean A1 length (1.75 ± 0.10 mm, *P* = 0.011), the mean A2 length (1.69 ± 0.34 mm, *P* = 0.035), and the mean PD (3.41 ± 0.92 mm, *P* < 0.001) were significantly lower in eyes with POAG. Conversely, the mean A1 velocity was significantly higher in eyes with POAG (0.16 ± 0.01 m/s) than in the controls (0.14 ± 0.01 m/s, *P* < 0.001). The mean CCR was significantly higher in the POAG group (7.25 ± 0.61 mm) than in the control eyes (6.59 ± 0.63 mm, *P* = 0.035). All the other analyzed parameters showed no significant difference.

**Table 1:** The descriptive analyses of the demographic variables and Corvis ST parameters and the comparisons between the POAG and control groups.

|  | Healthy controls | POAG patients | Total | P |
| --- | --- | --- | --- | --- |
| n | 16 | 33 | 49 |  |
| Age, year | 42.2 (20.9) | 58.6 (7.8) | 53.2 (15.4) | <0.001 |
| Sex, n male (%) | 5 (31.3%) | 25 (75.8%) | 30 (61.2%) | 0.003 |
| Axial length, mm | 27.2 (1.4) | 26.9 (0.8) | 27.0 (1.0) | 0.542 |
| Corneal curvature, mm | 7.7 (0.2) | 7.8 (0.2) | 7.8 (0.2) | 0.137 |
| CCT, μm | 556.5 (36.1) | 528.6 (29.3) | 537.7 (34.0) | 0.003 |
| A1 time, ms | 7.41 (0.28) | 7.14 (0.23) | 7.23 (0.28) | 0.004 |
| A1 length, mm | 1.77 (0.16) | 1.74 (0.05) | 1.75 (0.10) | 0.011 |
| A1 velocity, m/s | 0.14 (0.02) | 0.16 (0.01) | 0.15 (0.02) | <0.001 |
| A2 time, ms | 21.77 (0.42) | 21.93 (0.37) | 21.88 (0.39) | 0.076 |
| A2 length, mm | 1.78 (0.41) | 1.65 (0.30) | 1.69 (0.34) | 0.035 |
| A2 velocity, m/s | -0.42 (0.08) | -0.43 (0.07) | -0.42 (0.08) | 0.532 |
| HC time, ms | 16.60 (0.37) | 16.84 (0.54) | 16.76 (0.50) | 0.134 |
| Def. amp., mm | 1.09 (0.10) | 1.16 (0.10) | 1.14 (0.10) | 0.088 |
| Peak distance, mm | 4.21 (0.66) | 3.02 (0.77) | 3.41 (0.92) | <0.001 |
| CCR, mm | 6.59 (0.63) | 7.25 (0.61) | 7.03 (0.69) | 0.035 |

CCR: Concavity curvature radius, HC: Highest concavity, DA:
Deformation amplitude, AL: Axial length, CCT: Central corneal thickness,
POAG: Primary open-angle glaucoma

The potential corneal biomechanical indicators of POAG in high myopia are analyzed in Table 2. Of the 10 analyzed Corvis ST parameters, four showed a significant association with glaucoma. After adjusting for age and sex, the odds ratios per standard deviation (SD) increase in the parameter were as follows: the A1 time was 0.25 (95% confidence interval [CI]: 0.08 to 0.75, *P* = 0.014), the A1 velocity was 6.09 (95% CI: 1.84 to 20.16, *P* = 0.003), the A1 length was 0.27 (95% CI: 0.07 to 0.96, *P* = 0.043), and the PD was 0.22 (95% CI: 0.08 to 0.61, *P* = 0.004).

**Table 2:** The logistic models of the associations between POAG and the Corvis ST biomechanical parameters in high myopes and the corresponding receiver operating characteristic (ROC) analyses (AUC=area under curve)

|  | Logistic models |  | ROC curves |  |  |  |
| --- | --- | --- | --- | --- | --- | --- |
|  | Odds Ratios | P | AUC (95% CI) | Cutoff points | Sensitivity | Specificity |
| A1 time | 0.25 (0.08, 0.75) | 0.014* | 0.77 (0.63, 0.92) | 7.35 ms | 0.85 | 0.62 |
| A1 velocity | 6.09 (1.84, 20.16) | 0.003* | 0.78 (0.62, 0.94) | 0.157 m/s | 0.76 | 0.81 |
| A1 length | 0.27 (0.07, 0.96) | 0.043* | 0.52 (0.30, 0.74) | 1.73 mm | 0.79 | 0.56 |
| A2 time | 2.23 (0.89, 5.58) | 0.086 | - | - | - | - |
| A2 length | 0.41 (0.16, 1.09) | 0.075 | - | - | - | - |
| A2 velocity | 0.84 (0.4, 1.77) | 0.644 | - | - | - | - |
| HC time | 1.73 (0.78, 3.85) | 0.178 | - | - | - | - |
| Def. amp. | 2.02 (0.87, 4.73) | 0.103 | - | - | - | - |
| Peak distance | 0.22 (0.08, 0.61) | 0.004* | 0.87 (0.77, 0.97) | 3.26 mm | 0.73 | 1.00 |
| CCR, mm | 2.34 (0.91, 6.02) | 0.077 | - | - | - | - |

ORs: Odds ratios, AUC: Area under the curve, CI: Confidence interval, ROC: Receiver operating characteristic, CCR: Concavity curvature radius, HC: Highest concavity, DA: Deformation amplitude. \* $P < 0.05$

The four biomechanical parameters that were significantly associated with glaucoma in the logistic models were further analyzed with the ROC curves in Table 2. The Corvis ST parameter with the largest area under the curve (AUC) was PD, which was at 0.87 (95% CI: 0.77 to 0.97); the cutoff point for diagnosis was 3.26 mm. The A1 time and the A1 velocity had similar AUC, which were at 0.77 (95% CI: 0.63 to 0.92) and 0.78 (95% CI: 0.62 to 0.94), respectively. For A1 length, although it was significantly associated with the state of POAG in the logistic model (*P* = 0.043), the AUC was only 0.52 with a 95% CI containing 0.5 (95% CI: 0.30 to 0.74), indicating that it was not a good indicator of POAG in high myopia. Figure 1 shows the ROC curves of the four parameters, supporting the findings in Table 2.

**Figure 1:**
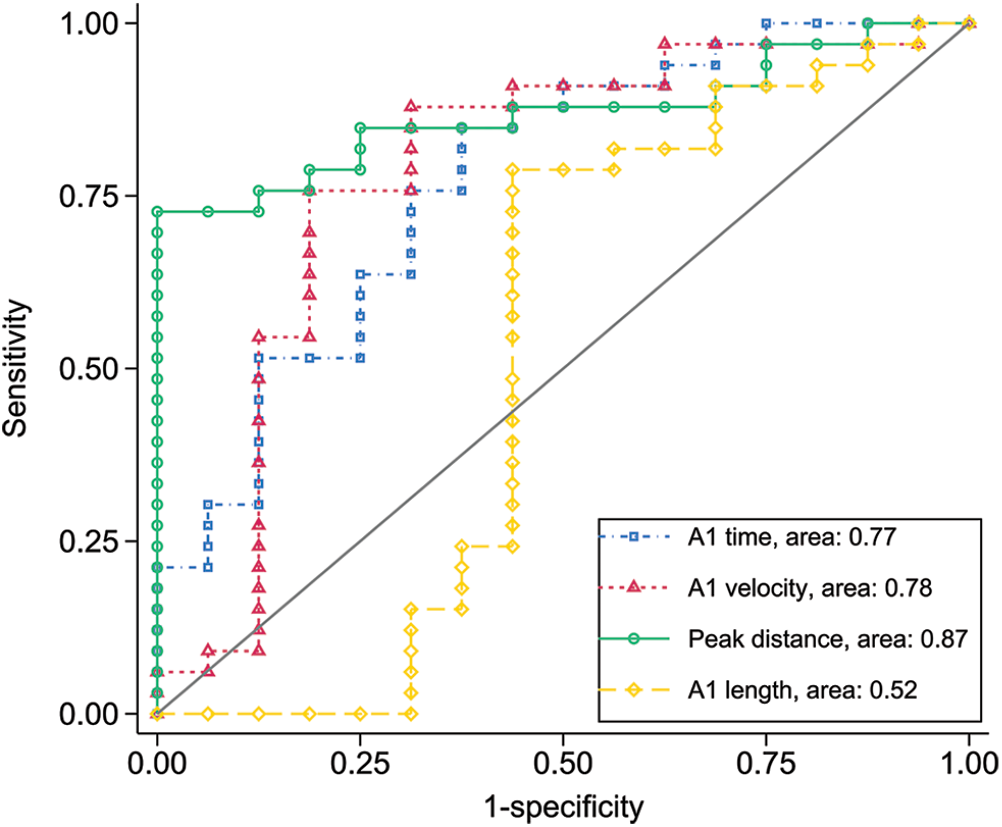
The receiver operating characteristic curves of four Corvis ST parameters, the peak distance, A1 time, A1 length and, A1 velocity

After analyzing the diagnostic value of each parameter using the ROC curve, a multivariate model (linear SVM) was built with the three candidate predictors, A1 time, A1 velocity, and PD. The fitting separating hyperplane was 1.97 × A1 time − 0.17 × A1 velocity + PD − 18.2 = 0. Considering the coefficient of A1 velocity was small (−0.17), A1 velocity was removed from the model, and a linear SVM model incorporating A1 time and PD was built. As shown in Figure 2, the separating line was 1.97 × A1 time + PD − 18.26 = 0, which effectively distinguished between the two groups, and the classification accuracy was 0.898 (=44/49) in the sample.

**Figure 2:**
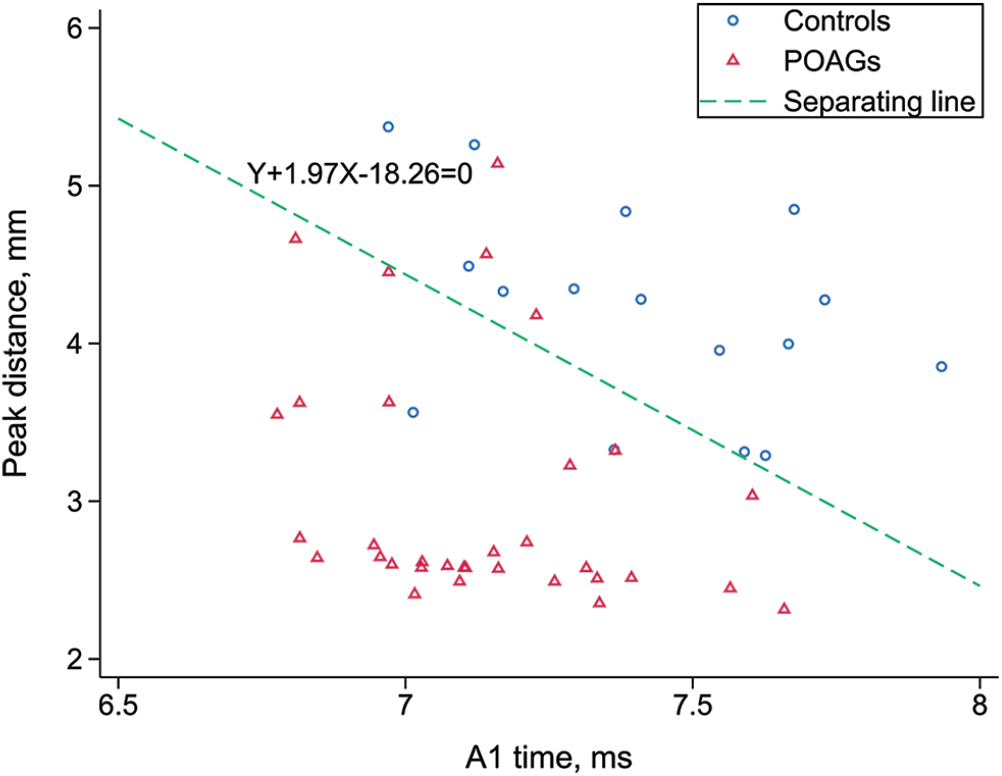
Plot of the separating hyperplane of the linear support vector machine model. POAG: Primary open-angle glaucoma

The sample size was analyzed based on the linear diagnostic models in this study (logistic and linear SVM). The events per predictor (EPP) was calculated, which was 33 events/3 predictors = 11 in this study, which satisfied the rule of minimum EPP at 10.^[12]^

## Discussion

This study focused on the challenging problem of diagnosing glaucoma in high myopia from a new perspective: corneal biomechanics. Table 1 shows a significantly smaller A1 time, A1 length, A2 length, PD, and higher A1 velocity in eyes with POAG. This indicated that among the high myopes, the cornea of eyes with POAG was stiffer than the normal eyes. The findings were supported by previous publications.^[13,14]^ A stiffer cornea in glaucoma has been reported since the era of the ocular response analyzer,^[15]^ which was reported as a risk factor independent of IOP.^[9]^ This demonstrated that among high myopes, corneal stiffness was significantly greater in POAG patients than in healthy controls.

In this study, A1 time, A1 velocity, A1 length, and PD were significantly associated with POAG, indicating that these four parameters may serve as diagnostic indicators in high myopia. In the ROC analyses, the AUC of PD was the largest, making it the most effective predictor in this study, followed by two parameters with similar AUCs: A1 velocity and A1 time. The diagnostic values were evaluated based on the AUCs, which were acceptable for A1 velocity and A1 time (between 0.7 and 0.8) and excellent for PD (over 0.8).^[16]^ According to previous publications, the diagnostic value of Corvis ST parameters has been indicated in various types of glaucoma,^[17,18]^ supporting our findings in this study.

The separating line of the linear SVM was then fitted as 1.97 × A1 time + PD − 18.26 = 0, based on which we proposed a *Q*-value that *Q* = 1.97 × A1 time (ms) + PD (mm) − 18.26. As shown in Figure 2, if the *Q* is <0, the eye is classified as having POAG; otherwise, if the *Q* is >0, it is classified as control. Considering the SD of *Q* is 1.1, the value of *Q* could be roughly interpreted as the number of SDs from the separating line. On the other hand, the cases close to the borderline, or where *Q* value was close to 0, may be more prone to misclassification. The AUC of the *Q* value was 0.94 (95% CI: 0.88 to 1.0), indicating excellent diagnostic value in the sample.^[16]^ Considering only two predictors were included in the SVM model, the risk of overfitting is low.

Strategic implementation is essential to overcome the current accessibility constraints of Corvis ST. We propose a hub-and-spoke model where tertiary centers conduct biomechanical phenotyping on high-risk high myopes with discordant conventional glaucoma test results. Risk scores derived from these assessments will guide community clinics in implementing personalized monitoring intervals (e.g., 6- vs. 12-month follow-ups), optimizing resource utilization while leveraging biomechanical insights.

## Conclusion

We found that Corvis ST parameters were significantly different between eyes with POAG and the control eyes in high myopes, and the direction of the changes indicated a stiffer alteration of the cornea in patients with POAG. The logistic and ROC analyses indicated several Corvis ST parameters as potential disease indicators. The SVM model showed a strong ability to differentiate between the POAG and control groups in the sample. This study suggests the Corvis ST parameters as potential biomechanical biomarkers in the diagnosis of POAG among high myopes.

## Data Availability

All data produced in the present study are available upon reasonable request to the authors

